# Seroprevalence of anti-SARS-CoV-2 IgG antibodies in children with household exposition to adults with COVID-19: preliminary findings

**DOI:** 10.1101/2020.08.10.20169912

**Authors:** Danilo Buonsenso, Piero Valentini, Cristina De Rose, Davide Pata, Dario Sinatti, Domenico Speziale, Rosalba Ricci, Angelo Carfì, Francesco Landi, Maurizio Sanguinetti, Michela Sali, the Gemelli Against COVID-19 Post-Acute Care Study Group

## Abstract

Wheather children are easily susceptible to SARS-CoV-2 infection is still a debated question and a currently a hot topic, particularly in view of important decisions on school opening. For this reason, we decide to describe preliminary data showing the prevalence of anti-SARS-CoV-2 IgG in children with known household exposure to SARS-CoV-2.

Our report shows that household transmission of SARS-CoV-2 is high in both adults and children, with similar rates of SARS-CoV-2 IgG in all age groups, including the younger children. A total of 44 out of 80 household contacts (55%) of index patients had anti SARS-CoV-2 IgG. In particular, 16 (59,26%) adult partners had IgG antibodies compared with 28 (52,83%) of pediatric contacts (P > 0.05). Among the pediatric population, children ≥ 5 years of age had similar probability of having SARS-CoV-2 IgG (21/39, 53.8%) compared with those < 5 years (7/14, 50%) (P > 0.05). Adult partners and children also had a probability of having SARS-CoV-2 IgG. Interestingly, 35.7% of children and 33.3% of adults with SARS-CoV-2 IgG were previously diagnosed as COVID-19 cases.

Since this evidence of high rate of IgG in children exposed to SARS-CoV-2 has public health implication, with this comment we highlight the need of establishing appropriate guidelines for school opening and other social activities related to childhood.

---

Eight months after the first outbreak in China, the SARS-CoV-2 pandemic is continuing to put a strain on the health systems around the world. A much-debated aspect of the disease concerns its impact on children.

The study of the prevalence and contagiousness of the disease among children produced conflicting results: on the one hand, low infection rates among children (1) and a low number of serious diseases (2) have been described, on the other hand, a serious, albeit rare, inflammatory complication has been reported (3). Similarly, on the one hand, low infectiousness rates in the school environment have been described (4), while a recent published data highlighted an easy spread of the disease among children in a summer camp (5). This is of particular concern as the return to school is one of the most critical steps to be addressed in phase two of the pandemic.

To better characterize the possibility of children to be infected with the novel coronavirus, we studied the prevalence of anti-SARS-CoV-2 IgG in children with known household exposure to SARS-CoV-2, and to compare this data with adult partners exposed to the same index case (supplementary material for more methodological details) (6, 7). The full study is still ongoing, aiming to evaluate the presence of neutralizing antibodies in adults and children exposed to SARS-CoV-2 and to prospectively follow-up these families in order to understand if a specific antibody pattern protects from further infection.

Of 405 adults with COVID-19 evaluated in the outpatient post COVID unit, 33 were living with children younger than 18 years of age. Of those, 30 (90.9%) agreed to participate and were considered index cases. At the time of COVID-19 diagnosis of the index case, a total of 80 household contacts were living in the same household and were enrolled in the study of which 53 were children and 27 adult partners (figure 1).

Anti SARS-CoV-2 IgG were present in 44 (55%) household contacts of which 16 (59,26%) were adult partners and 28 (52,83%) were pediatric contacts (P > 0.05). Similar relative frequencies of seropositivity where present in children ≥ 5 years of age (21/39, 53.8%) and those < 5 years (7/14, 50%) (P > 0.05). Adult partners and children also had a similar relative frequency of having SARS-CoV-2 IgG. Interestingly, ten children (35.7%) and five adults (33.3%) adults with SARS-CoV-2 IgG reported a history of diagnosis of COVID-19.

Our report shows that household transmission of SARS-CoV-2 is high in both adults and children, with similar rates of SARS-CoV-2 IgG in all age groups, including the younger children. SARS-CoV-2 infection rate was higher in our cohort compared with a large contact tracing study performed in South Korea (8). This difference could be related to the study design involving serology. Importantly, more of the 60% of the household contacts we evaluated, including children, were not diagnosed SARS-CoV-2 infection before this study probably because most of them did not develop symptoms and were not tested with nasopharyngeal swabs. This strongly suggests that the real burden of the SARS-CoV-2 pandemic, in particular the pediatric cases, is hugely underestimated (9, 10).

Our findings have public health implications, since highlight that children of all age groups have high probability of being infected with SARS-CoV-2 if closely exposed to an index case. Considering that recent studies suggested that children can have similar (or even higher) viral loads than adults on nasopharyngeal swabs, even if asymptomatic (11), our study further supports the importance of the need of appropriate procedural guidelines of childhood activities, including schools, in the time of COVID-19. Although access to school is a priority right of children with several benefits on all aspects of child development, the growing evidences that children can easily being infected with SARS-CoV-2 and contribute to virus spread should be used to implement public health recommendations, including hand and respiratory hygiene, physical distancing, masking and active surveillance to reduce and/or prevent of SARS-CoV-2 transmission within childhood environments.

## Data Availability

available upon request

## Data availability

All the data and material are available upon request to the corresponding author.

### Conflict of interest

None of the participants has any conflict of interest to declare.

## Acknowledgments

the members Gemelli Against COVID-19 Post-Acute Care team are listed in the supplementary material

---

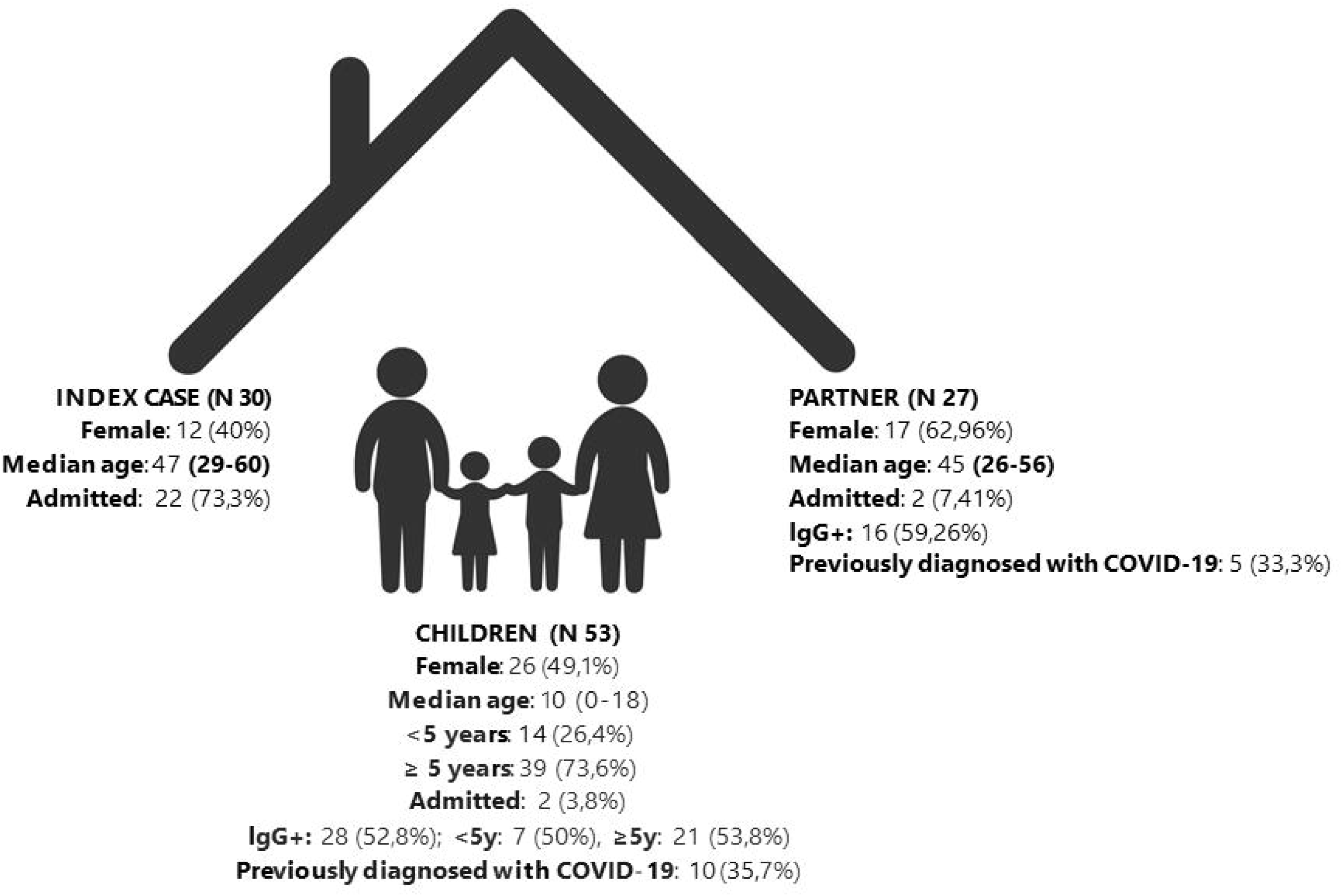

## Notes

### Competing Interest Statement

The authors have declared no competing interest.

### Clinical Trial

not registered for organizational procedures

### Funding Statement

nothing to declare

### Author Declarations

The study has been approved by the Ethic Committee of the Fondazione Policilnico Universitario A. Gemelli IRCCS, Rome, italy (ID 3078)

